# Refining “long-COVID” by a prospective multimodal evaluation of patients with long-term symptoms related to SARS-CoV-2 infection

**DOI:** 10.1101/2021.04.08.21255167

**Authors:** Marc Scherlinger, Renaud Felten, Floriane Gallais, Charlotte Nazon, Emmanuel Chatelus, Luc Pijnenburg, Amaury Mengin, Adrien Gras, Pierre Vidailhet, Rachel Arnould-Michel, Sabrina Bibi-Triki, Raphaël Carapito, Sophie Trouillet-Assant, Magali Perret, Alexandre Belot, Seiamak Bahram, Laurent Arnaud, Jacques-Eric Gottenberg, Samira Fafi-Kremer, Jean Sibilia

## Abstract

**Background:** COVID-19 long-haulers or “long-COVID” represent 10% of COVID-19 patients and remain understudied.

**Methods:** In this prospective study, we recruited 30 consecutive patients seeking medical help for persistent symptoms (> 30 days) attributed to COVID-19. All reported a viral illness compatible with COVID-19. The patients underwent a multi-modal evaluation including clinical, psychological, virological, specific immunological assays and were followed longitudinally.

**Results:** The median age was 40 [interquartile range: 35-54] and 18 (60%) were female. After a median time of 152 [102-164] days after symptom onset, fever, cough and dyspnea were less frequently reported as compared with the initial presentation, but paresthesia and burning pain emerged in 18 (60%) and 13 (43%) patients, respectively. The clinical examination was unremarkable in all patients although the median fatigue and pain visual analogic scales were 7 [5-8] and 5 [2-6], respectively.

Extensive biological studies were unremarkable, as were multiplex cytokine and ultra-sensitive interferon-a2 measurements. At this time, nasopharyngeal swab and stool RT-PCR were negative for all tested patients. Using SARS-CoV-2 serology and IFN-γ ELISPOT, we found evidence of a previous SARS-CoV-2 infection in 50% (15/30) of patients, with objective evidence of lack or waning of immune response in two. Finally, psychiatric evaluation showed that 11 (36.7%), 13 (43.3%) and 9 (30%) patients had a positive screening for anxiety, depression and post-traumatic stress disorder, respectively.

**Conclusions:** Half of patients seeking medical help for long-COVID lack SARS-CoV-2 immunity. The presence of SARS-CoV-2 immunity did not cluster clinically or biologically long haulers, who reported severe fatigue, altered quality of life, and exhibited psychological distress.

**Key points:**

- Among 30 consecutive patients reporting persistent symptoms (median 6 months) self-attributed to COVID-19, pain, fatigue and disability were reported in virtually all patients.
- More than one third of patients suffer from psychological disorders such as anxiety, depression and/or post-traumatic stress disorder, regardless of SARS-CoV-2 immunity.
- At the time of evaluation, only 50% of patients had cellular and/or humoral sign of a past SARS-CoV-2, and serology positivity varied depending of the kit used.
- Exhaustive clinical, biological and immunological evaluations failed to find an alternative diagnosis, or to identify specific cytokine signature including type I interferon.

## Introduction

After SARS-CoV-2 infection, some patients may continue to experience symptoms for months (1), a condition called “long-COVID” or “COVID-19 long-haulers” (2). These long-term symptoms contribute to the worldwide COVID-19 burden and have been extensively communicated by the patients and some physicians (3,4). Long-COVID manifestations are clinically diverse, and their underlying mechanisms are likely to be multiple. Patients who underwent long hospitalization because of severe disease may exhibit lung or heart chronic injury due to the severe immune response (5,6), micro- or macrovascular thrombotic neurological complications (7), and/or physical deconditioning (8). Indeed, a recent cohort study found that among 1733 hospitalized COVID-19 patients, 76% reported at least one symptom after 6 months, and objective pulmonary abnormalities were found in more than 20% (9). However, these pathological mechanisms are less likely to explain the manifestations in patients with initially non-severe COVID-19 who experience long-lasting symptoms. Although 10% of COVID-19 patients may show chronic symptoms (>12 weeks) (10), only scarce data are available on the clinical presentation, biological characteristics and overall prognosis of such patients (11,12). Moreover, the virological definition of the long-COVID entity in terms of serology and other specific immunological assays is still not precisely established (11,13).

The aim of our study was to refine the clinical and biological characteristics of long-COVID by conducting a multimodal evaluation of consecutive patients seeking medical help for persistent symptoms attributed to COVID-19 (long-COVID).

## Methods

### Patients and ethical considerations

We included 34 consecutive patients seeking medical help for persistent symptoms attributed to COVID-19 for a systematic prospective study. Patients were recruited in the “Grand-Est” region of France, the area with the highest incidence of SARS-CoV-2 infection during the first epidemical wave in France (February to April 2020) (14). Information about a long-COVID consultation in our tertiary center was advertised through local media and social networks, and consulting patients were then asked to participate in the study. All patients provided written informed consent. Age-matched patients with a history of non-severe COVID-19 (positive humoral and cellular response) without persistent symptoms were controls for the immunological assays. This study is part of the “COVID-HUS” study, which was approved by the ethics committee of the University Hospital of Strasbourg (NCE–2020– 51).

### Medical evaluation

All patients were evaluated at our reference center for rare systemic and autoimmune diseases. Five physicians (MS, RF, EC, LP and JS) conducted the clinical evaluation with a physical examination and a standardized questionnaire on medical and SARS-CoV-2–related infection history. Dedicated questionnaires were used to evaluate subjective symptoms: DN4 for neuropathic pain (15), Fibromyalgia Rapid Screening Tool (FiRST (16)) and visual analog scales for fatigue/pain/dryness. Additionally, patients underwent systematic blood testing to rule out alternative diagnoses (full list in supplementary file 1).

### Virologic and immunologic evaluation

Blood samples were collected at the same time as the clinical evaluation for immuno-virologic assays. The testing included extensive serology evaluation with 4 commercial assays. One lateral flow assay tested for the receptor binding 2 (RBD) of the spike protein as an antigenic source (Biosynex BSS IgM/IgG assay), and two enzyme-linked immunosorbent assays (ELISA) targeted the RBD (Wantai total antibody) and the S1 domain (Euroimmun IgG). Finally, ELISA was used to detect anti-nucleocapsid (anti-N) IgG (Abbott Architect IgG). To explore the SARS-CoV-2–specific T-cell response, heparin-anticoagulated blood tubes were collected for interferon gamma (IFN-γ) enzyme-linked immunospot (ELISPOT) assay. Peripheral blood mononuclear cells were isolated by using Ficoll-Paque gradient, and CD3^+^ cell frequencies were measured by flow cytometry. Anti–IFN-γ antibody-coated wells (UCytech) were seeded with 200,000 CD3^+^ cells per well. Cells were stimulated in duplicate with overlapping 15-mer peptide pools spanning SARS-CoV-2 structural proteins (spike protein [S1 and S2], nucleoprotein, membrane and envelope proteins) and accessory proteins (NS3, NS7A, NS8, NS9B) as well as the spike glycoprotein of human coronaviruses 229E (ES1, ES2) and OC43 (OS1, OS2) (PepMix, JPT Peptide Technologies). Culture medium was a negative control and phytohemagglutinin (PHA) a positive control. Peripheral blood mononuclear cells were cultured overnight (20 ± 4 hr) before enzymatic revelation of IFN-□ capture (UCytech). Spots were counted by using an ELISPOT reader (AID) and results are expressed as mean number of spot-forming units/10^6^ CD3^+^ cells after subtraction of the background value. Positivity was defined as a T-cell response ≥ 3 times the standard deviation of the negative controls. To complete the immunological evaluation, IFN-*α*2 (with Single Molecule Array; SIMOA, Quanterix (17)) and cytokines (with a custom multiplex cytokine assay; Luminex Thermo Fisher) were measured from serum samples.

### Psychological evaluation

A standardized interview was conducted by a clinical psychologist (R.A-M). The interview aimed at evaluating psychiatric history and treatments, presence of sleep issues and increase of toxic or anxiolytic use. The impact of the COVID-19 pandemic on quality of life was assessed by using the Medical Outcomes Study 36-item Short-form (SF-36) and Health Assessment Questionnaire (HAQ). Patients’ perspectives on the pandemic crisis were assessed. Finally, validated translated questionnaires were used to screen for anxiety/depression (Hospital Anxiety and Depression Scale [HADS-A/D]) and post-traumatic stress disorder (PTSD checklist [PCL-5] for the Diagnostic and Statistical Manual of Mental Disorders, Fifth edition [DSM-5]). Positive screening for these mental disorders was confirmed with a score ≥ 10 for HADS-Anxiety, ≥7 for HADS-Depression (18), and ≥ 31 for PCL-5 (19).

### Statistics

Quantitative data are reported with median (interquartile range [IQR]) and were compared by non-parametric Mann-Whitney test. Categorical data are reported with number (%) and were compared by chi-square or Fisher exact test, as appropriate. To compare initial and persistent clinical features, McNemar’s test was used with Bonferroni’s correction for multiple testing. Statistical analyses involved using GraphPad Prism 7.0. P < 0.05 was considered statistically significant.

## Results

### Patients

A total of 34 consecutive patients seeking help for persistent symptoms attributed to COVID-19 visited to our center between June and August 2020; 30 were included in the study. Reasons for non-inclusion were refusal of physical consultation because of geographical distance (n=3), spontaneous improvement of the condition (n=1). In total, 60% were women (18/30) and the median age was 40 years (IQR 35-54). The cohort included 2 married couples, and other patients were unrelated. Before the initial presentation, none of the patient reported chronic pain nor the use of analgesics. Other characteristics are in **Table 1**.

**Table 1:** **Characteristics of the whole population and patients with or without cellular and/or humoral immunization**.

| Characteristics | Total population (n = 30) | Immunized (n = 15) | Non-immunized (n = 15) | p-value |
| --- | --- | --- | --- | --- |
| **Demographics** |  |  |  |  |
| Age (median, IQR) | 40 [35-54] | 40 [31-58] | 39 [35-45] | *ns* |
| Female sex | 60% (18) | 46.7% (7) | 73.3 (11) | *0.14* |
| Close contact with confirmed COVID-19 patients | 43.3% (13/30) | 46.7% (7/15) | 40% (6/15) | *ns* |
| **Risk factors for severe SARS-CoV-2 infection (history of)** |  |  |  |  |
| BMI (kg/m<sup>2</sup>), (median, IQR) | 22.6 [20.4-26.3] | 23.6 [21.2-27.8] | 22.4 [19.6-24.5] | *ns* |
| > 25 | 30% (9) | 40% | 20% |  |
| > 30 | 20% (6) | 26.7% | 13.3% |  |
| Diabetes | 10% (3) | 13.3% (2) | 6.7% (1) | *ns* |
| Hypertension | 3.3% (1) | 6.7% (1) | 0% | *ns* |
| Myocardial infarction | 0% |  |  | *ns* |
| Cerebrovascular event | 6.7% (1) | 13.3% (2) | 0% | *ns* |
| Respiratory disease | 0% |  |  | *ns* |
| Renal failure | 0% |  |  | *ns* |
| Liver failure | 0% |  |  | *ns* |
| Cancer | 3.3% (1) | 6.7% (1) | 0% | *ns* |
| Smoking |  |  |  |  |
| History | 43.3% (13) | 53.3% (8) | 33.3% (5) | *ns* |
| Active | 3.3% (1) | 0% | 6.7% (1) |  |
| **Socio-economic** |  |  |  |  |
| Married | 28.6% | 30.8% | 26.7% | *ns* |
| Working status |  |  |  |  |
| Currently working | 83.3% | 66.7% | 100% | *ns* |
| Unemployed | 10% | 20% | 0% |  |
| Retired | 6.67% | 13.3% | 0% |  |
| Education level |  |  |  |  |
| Below high school | 6.7% | 13.3% | 0% | *ns* |
| High school | 16.7% | 26.7% | 6.7% |  |
| ≤3 years post-high school | 26.7% | 13.3% | 40% |  |
| College graduate | 50% | 46.7% | 53.3% |  |

BMI, body mass index; ns, not significant

### Initial presentation

All patients declared a viral illness compatible with COVID-19. Initial symptoms occurred between 1 February and 9 April 2020 and were mainly characterized by fever (60%), myalgia (76.7%), cough (73.3%) and anosmia (43.3%) (**Figure 1**). Among 9 patients who underwent nasopharyngeal SARS-CoV-2 RT-PCR within 5 weeks after symptom onset, 55.6% (5/9) had a positive result and all RT-PCR tests conducted after this time were negative (n = 11). Seven patients consulted in the emergency department, and one was hospitalized in a conventional hospital unit to receive oxygen for 7 days, without specific treatment. All other patients were cared for at home. A total of 13 patients were prescribed treatments: 2 received hydroxychloroquine 400 mg/day, 4 prednisone 1 mg/kg for 5-7 days, 8 antibiotics (azithromycin, n = 6; amoxicillin, n = 2) and 2 low-dose aspirin. The other 17 patients received only symptomatic treatment (acetaminophen).

**Figure 1:**
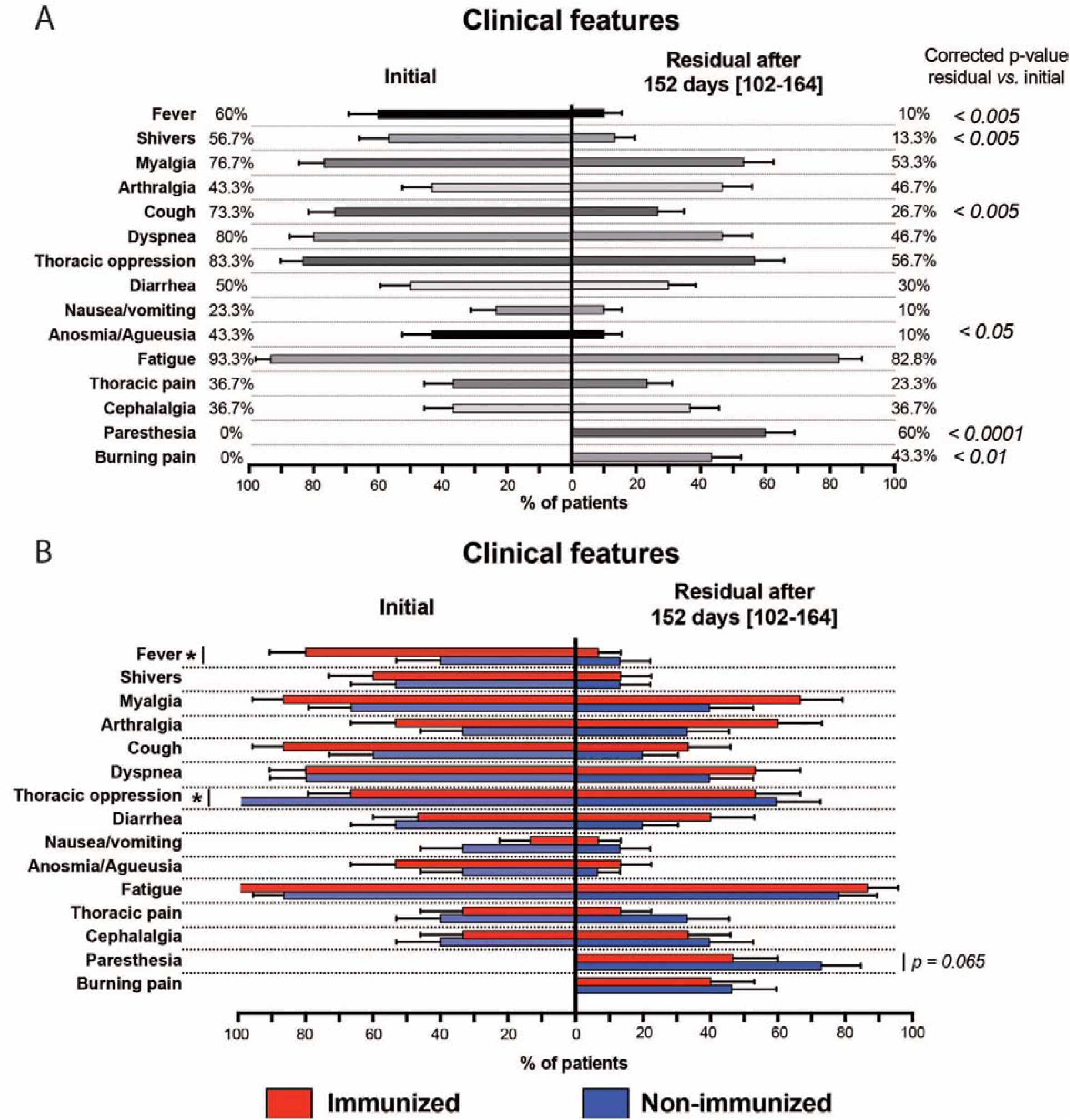
Initial and residual clinical features of patients with persistent symptoms self-attributed to COVID-19. (A) Residual symptoms collected at a median of 152 days [interquartile range (IQR) 102-164] after initial presentation (n = 30 patients). Columns indicate symptom prevalence and bars are mean± standard error measure. P-values were calculated with McNemar’s test with Bonferroni correction for multiple analysis. (B) Comparison of initial/residual symptoms between immunized and non-immunized patients. Columns indicate symptom prevalence in immunized (defined by ELISA or serology; red) and non-immunized patients; bars are mean± standard error measure. *p< 0.05 by chi-square or Fisher’s exact test (if appropriate).

### Persistent clinical features

Patients were clinically evaluated after a median of 152 days (IQR 102-164) after the reported initial symptom onset. Seventeen (56.7%) reported a resolution of initial symptoms after a median of 21 days (IQR 15-33), followed by a resurgence after a median of 21 days later (IQR 15-44). Conversely, the 13 other patients had no free-symptom intervals. Persistent symptoms had a cyclical pattern in 28 (93.3%) patients and were mostly represented by fatigue, myalgia and thoracic oppression (**Figure 1A**). Fever, shivering and cough were significantly less frequent as compared with the initial presentation (p < 0.005 for all; **Figure 1A**). Fatigue was severe for most patients and rated at a median of 7 (IQR 5-8) on a 10-point scale, with pain rated at 5 (IQR 2-6).

Overall, 60% and 43.4% of patients exhibited diffuse paresthesia and burning pain later after the initial presentation (**Figure 1A**). The DN4 questionnaire screening neuropathic pain was positive (≥4/10) for 50% (15/30) of patients and the FiRST questionnaire screening for fibromyalgia-like symptoms was positive (≥ 5/6) for 56.6% (17/30; **supplementary figure 1**).

The clinical examination including neurological examination was unremarkable. At this time, no patients received corticosteroids, non-steroidal anti-inflammatory drugs or opioids. Finally, 16 (53.3%) patients reported a trend toward a decrease of symptoms over time.

### Virologic and specific immunologic evaluation

Specific analyses related to SARS-CoV-2 infection were conducted at a median of 174 days (IQR 144-215) after symptom onset. At this time, in 18 (60%) and 6 (20%) of patients underwent nasopharyngeal and stool RT-PCR for SARS-CoV-2; all tests were negative.

Exploration of the cellular immune response by SARS-CoV-2 IFN-γ ELISPOT assay revealed that 15 (50%) patients had a positive response to at least SARS-CoV-2 nucleocapsid and spike proteins (considered ELISPOT-positive, **Figure 2A**). Among ELISPOT-positive patients, 73.3% (11/15) also had a cellular response against non-structural SARS-CoV-2 as compared with only 1 (6.7%) of ELISPOT-negative patients (p < 0.0001). Cellular response to the spike protein of human coronavirus 229E and OC43 was similar in both SARS-CoV-2 ELISPOT groups (E+100% vs E– 86.7% and E+ 80% vs E– 73.3%).

**Figure 2:**
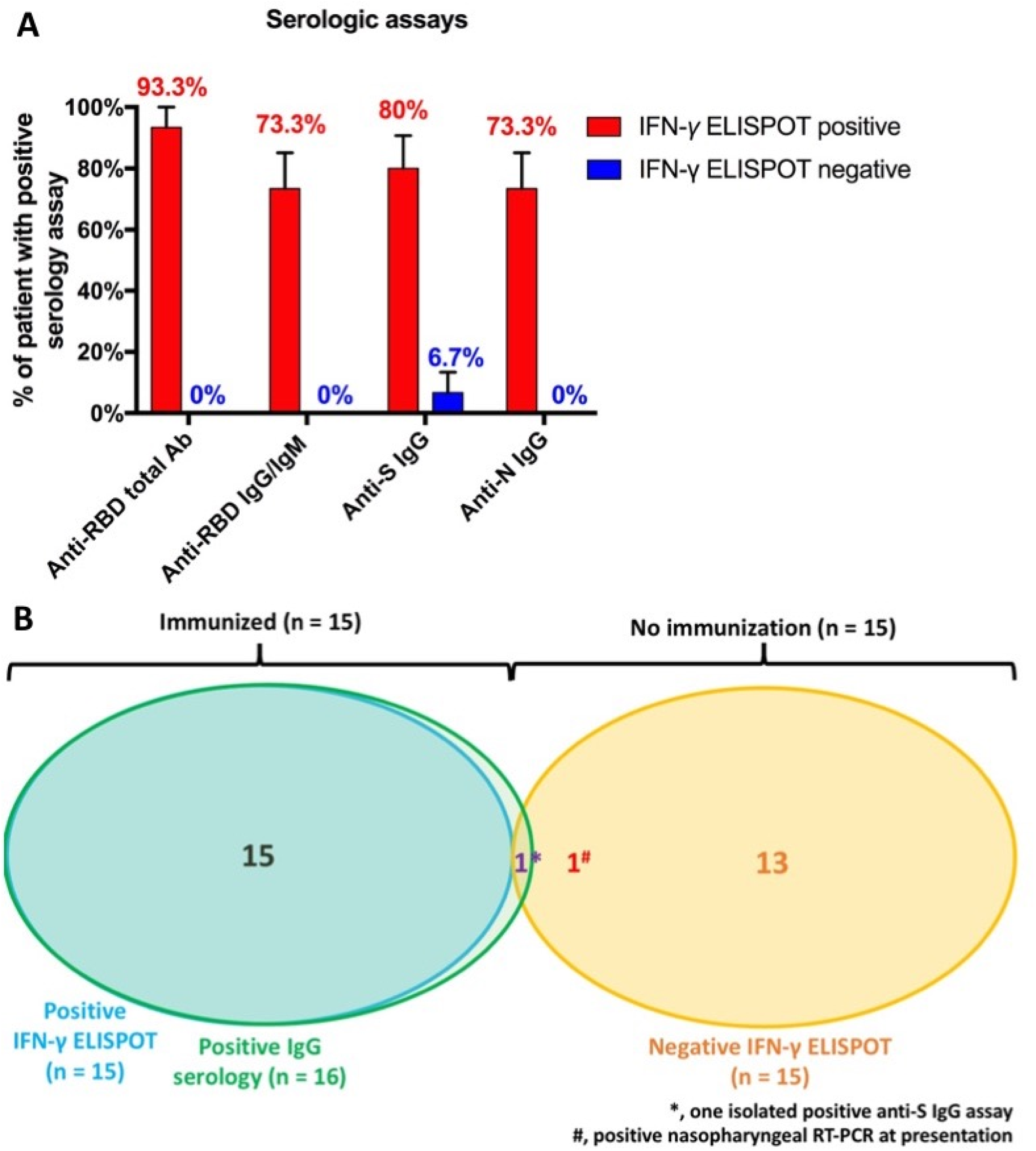
Specific immunological responses to SARS-CoV-2 in 30 patients reporting persistent symptoms self-attributed to long-COVID. *(A) Results of SARS-CoV-2 serologic assays based on the interferon-γ (IFN-γ) ELISPOT-results (n = 15/group). Anti-RBD total antibody (Wantai total antibody); anti-RBD IgG/IgM (Biosynex BSS IgM/IgG assay); anti-S IgG (Euroimmun); anti-N IgG (Abbott Architect). Data are mean± standard error measure prevalence of test positivity*. *(B) Two patterns of patients were identified: those with objective signs of SARS-CoV-2 immunity (cellular and humoral response, n = 15) and those without (n = 15). Positive IgG was defined as a positive result against spike, receptor binding domain or nucleocapsid protein*. *\*, One patient with virologically unproven initial presentation had an isolated anti-S IgG–positive assay result*. IFN-*γ* ELISPOT *and all other serological tests were negative*. *#, One with a virologically-proven initial presentation had negative SARS-CoV-2 ELISPOT (conducted at day 140) and serological assays (conducted at days 37, 85 and 140)*.

Among patients with a negative IFN-γ ELISPOT result, all but one had negative serology results (**Figure 2A**). This patient had an isolated anti-S IgG assay. Among the 15 patients with a positive IFN-γ ELISPOT result, all had at least one positive serological assay. In detail, 14 (93.3%) had a positive result for anti-RBD total antibodies, 11 (73.3%) anti-RBD IgG/IgM, 12 (80%) anti-S IgG and 11 (73.3%) anti-N IgG (**Figure 2A**). We dichotomized the patients into an immunized group (ELISPOT-positive and at least one positive serological assay, n = 15) and a non-immunzed group (ELISPOT-negative, n = 15; **Figure 2B**), with little difference in terms of symptoms (**Figure 1B**).

One ELISPOT-positive patient showed a decreasing signal for anti-N (equivocal to negative) and anti-S signal (positive to equivocal) between day 133 and day 251. Additionally, one patient with negative results for both IFN-γ ELISPOT and serology assays (conducted 140 days after symptom onset) previously had a positive nasopharyngeal RT-PCR SARS-CoV-2 result early after the first symptoms (**Figure 2B**). This patient also had 2 negative IgG serology tests for SARS-CoV-2 at 37 and 85 days after symptom onset (Biosynex ICT and CLIA Cobas Roche, respectively).

IFN-*α*2 levels were similar for patients with persistent symptoms (regardless of their immunity status) and those with a history of documented COVID-19 free of residual symptoms (sampled > 12 weeks after infection; **Figure 3**). Other cytokine levels were low or non-detectable and were similar between patients with and without immunization for SARS-CoV-2 (**Supplementary figure 2**).

**Figure 3:**
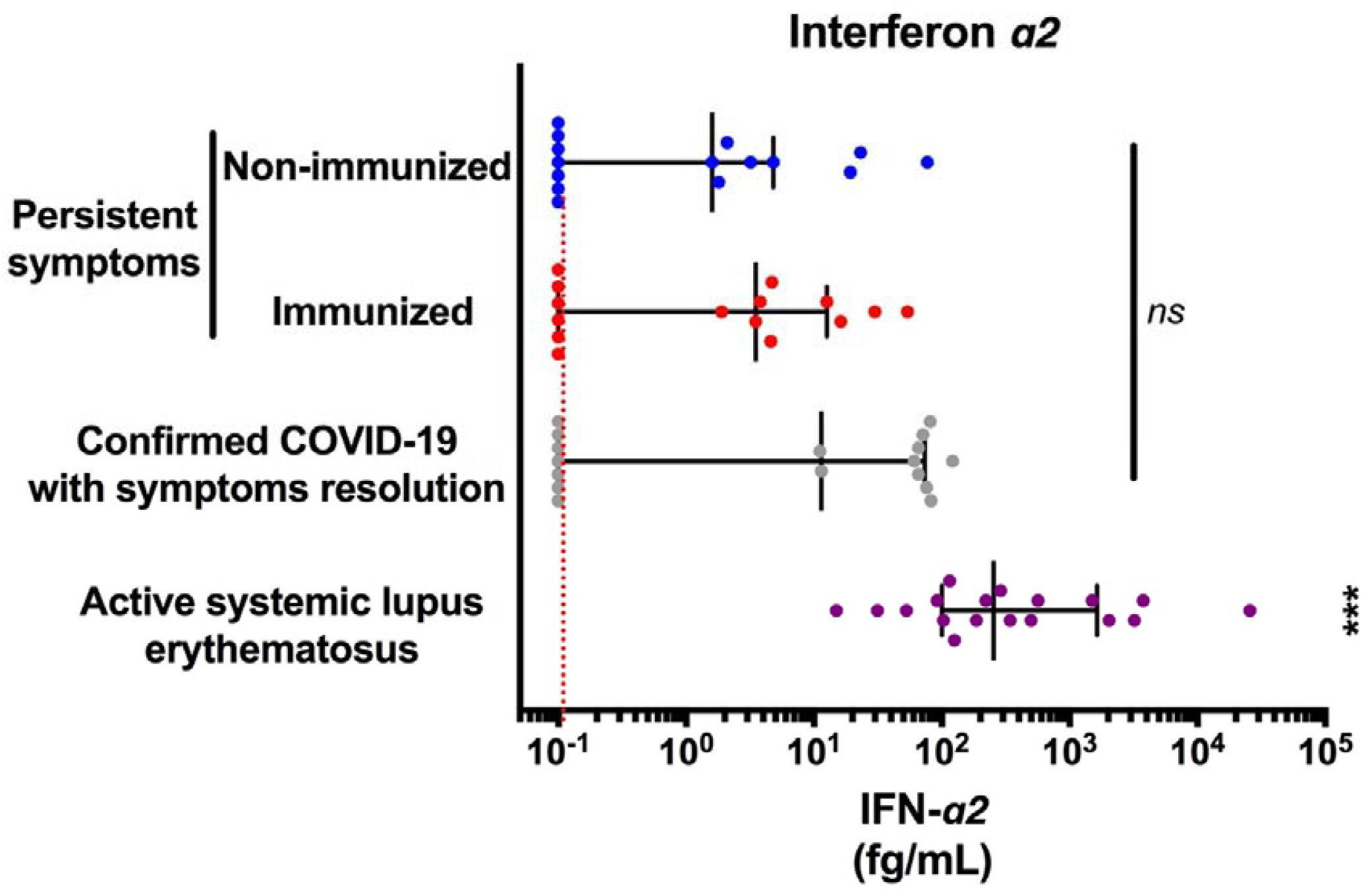
Normal levels of IFN-α2 for patients with persistent symptoms attributed to COVID-19. *Ultra-sensitive IFN-α levels were measured by using SIMOA in patients with persistent symptoms self-attributed to COVID-19. As a comparison, IFN-α2 levels of individuals with confirmed previous COVID-19 (serology and IFN-γ positive) without persistent* symptoms (*sampled at least 12 weeks after*; n = *17) and patients with active systemic lupus erythematosus (n = 18). Each point corresponds to a single patient; the central bar shows medians with interquartile ranges. The red dotted line shows the lower limit of detection. ns, non-significant; ***, p < 0*.*001 versus all other groups by non-parametric Kruskal-Wallis test with Dunn’s correction for multiple testing*.

### Biological evaluation

Biological analyses (**supplementary file 1**) were conducted at the same time as the clinical evaluation (152 days [IQR 102-164] after initial presentation). Routine biological test results were within normal limits for all but one patient (an iron-deficiency anemia that was further investigated and corrected). Values for markers of cardiac and muscle injury (troponin and creatine phosphokinase) and coagulopathy (D-dimers, fibrinogen) were normal, and serology for HIV, hepatitis C virus and Lyme disease was negative for all patients.

Screening for autoimmunity revealed low (1/160) and medium (1/320 to 1/640) titers of anti-nuclear antibodies in 12 and 3 patients, respectively. Low to medium anti-nuclear antibody titers were numerically more prevalent in SARS-CoV-2 immunized than non-immunized patients (66.7% vs 33.3%, p = 0.067). Screening for anti-extractable nuclear antigens, anti-double-stranded DNA, anti-citrullinated protein and anti-neutrophil cytoplasmic antibodies as well as rheumatoid factor was negative for all patients. Eight patients (4 in each immunization group) showed isolated low titers (< 3 times normal range) of anti-cardiolipin antibodies, with no history of thrombosis (IgM for 6, IgG for 2), and one patient was positive for lupus anticoagulant. After 12 weeks, the repeat antiphospholipid antibodies testing was negative for all patients.

### Psychological evaluation

The phone interview by a clinical psychologist took place after a median of 224 days [202-238] after initial symptom onset. In all, 10% (3/30) and 26.7% (8/30) of patients had a history of depression and anxiety disorders, respectively. Sleep issues were reported by 23 (73.3%) patients, and 4 (13.3%) had started an anxiolytic prescriptions (**Table 2**); 5 (16.7%) and 7 (23.3%) reported loss of employment and financial difficulties.

**Table 2:** **Psychological evaluation of the whole population and patients with and without cellular and/or humoral immunization**.

| **Psychological evaluation** | **Total population**<br>**(n =30)** | **Immunized**<br>**(n = 15)** | **Non-immunized**<br>**(n = 15)** | **p-value** |
| --- | --- | --- | --- | --- |
| **History** |  |  |  |  |
| Depression disorder | 10% (3) | 6.7% (1) | 13.3% (2) | *ns* |
| Anxiety disorder | 26.7% (8) | 26.7% (4) | 26.7% (4) | *ns* |
| **Context** |  |  |  |  |
| Loss of job due to pandemic | 16.7% (5) | 20% (3) | 13.3% (2) | *ns* |
| Financial difficulties | 23.3% (7) | 20% (3) | 26.7% (4) | *ns* |
| Death of close-ones due to COVID-19 | 0% |  |  |  |
| **Symptoms** |  |  |  |  |
| Sleep issues | 76.7% (23) | 80% (12) | 73.3% (11) | *ns* |
| Use of anxiolytics | 13.3% (4) | 20% (3) | 6.7% (1) | *ns* |
| Increased alcohol/tobacco use after diagnosis | 6.7% (2) | 13.3% (2) | 0% | *ns* |
| Probable anxiety disorder (HADS-A $\geq 10$) | 36.6% (11) | 33.3% (5) | 40% (6) | *ns* |
| Probable depression disorder (HADS-D $\geq 7$) | 43.3% (13) | 46.6% (7) | 40% (6) | *ns* |
| Probable post-traumatic stress syndrome (PCL-5 $\geq 31$) | 30% (9) | 33.3% (5) | 26.7% (4) | *ns* |
| **Patients' perception\***<br>**(N = 29)** |  |  |  |  |
| COVID-19 is dangerous | 79.3% (23) | 85.7 (12) | 73.3% (11) | *ns* |
| COVID-19 pandemic is worrisome | 41.4% (12) | 35.7% (5) | 46.7% (7) | *ns* |
| Support from family | 66.7% (20) | 73.3% (11) | 60% (9) | *ns* |
| Support from friends | 60% (18) | 73.3% (11) | 46.7% (7) | *ns* |
| Support from colleagues | 44% (11/25) | 36.4% (4/11) | 50% (7/14) | *ns* |
| Support from physicians | 20.7% (6) | 28.6% (4) | 13.3% (2) | *ns* |
| Good acknowledgment from the media | 10.3% (3) | 21.4% (3) | 0% | *0.10* |
| Good management of pandemics by the authorities | 20.7% (6) | 35.7% (5) | 6.7% (1) | *0.08* |

\*, answers were recorded on a 1-5 scale, and positivity was considered with score $\geq 4$ HADS, Hospital Anxiety and Depression Scale; A, anxiety; D, depression

HADS screening for anxiety and depression was positive for 11 (36.7%) and 13 (43.3%) of patients, respectively (**Table 2**). Using the PCL-5 questionnaire, 9 (30%) patients had scores compatible with PTSD (**Table 2**). These values did not differ by immunization status (**Figure** 4). Several components of the SF-36 scale, physical limitations, energy and pain, were severely affected, with no significant difference between patients immunized or not for SARS-CoV-2 (**supplementary figure 3**). Family, friends and colleagues were a major source of support for 20 (66.7%), 18 (60%) and 11/25 (44%) patients, respectively. Conversely, 7 (23.3%) patients felt significant support by physicians.

**Figure 4:**
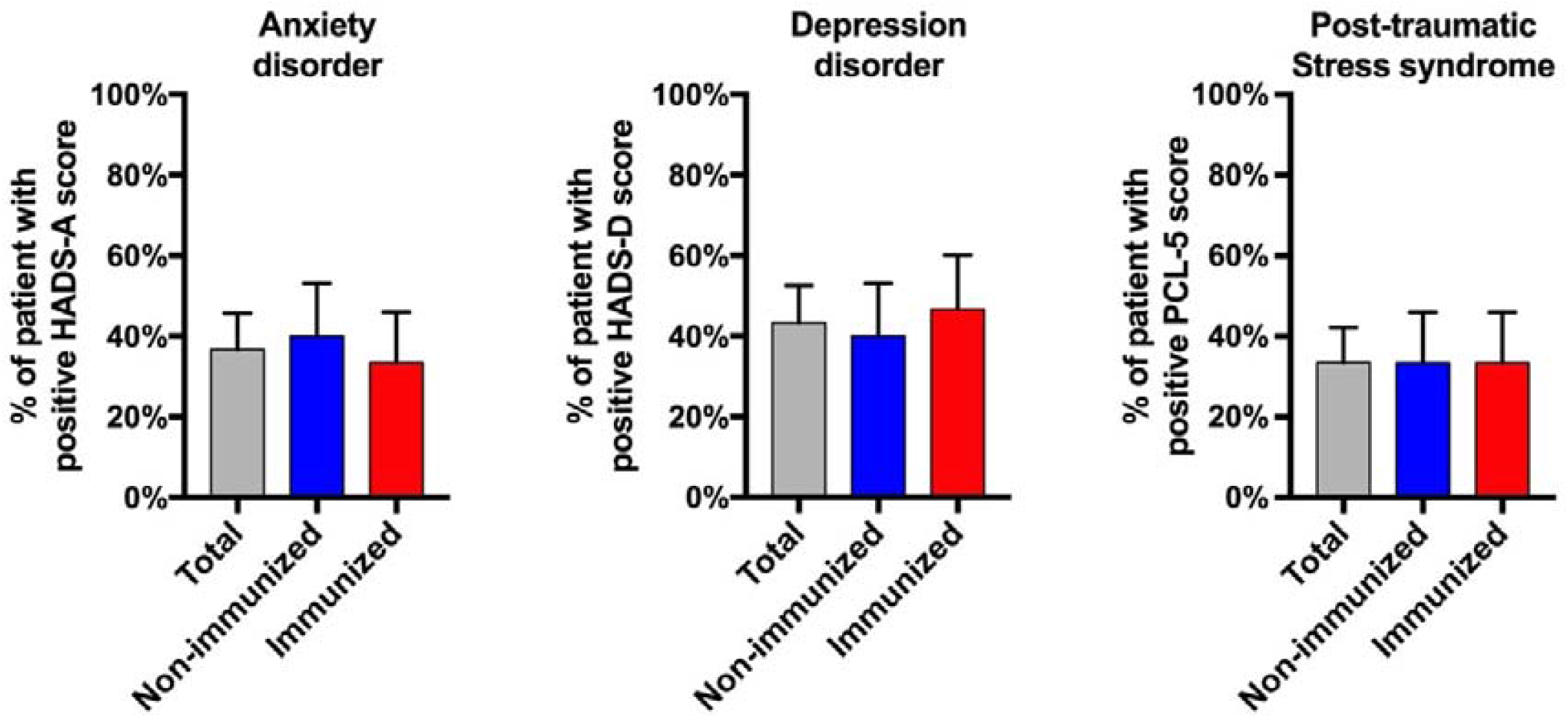
Prevalence of anxiety/depression disorders and post-traumatic stress syndrome in patients seeking medical help for persistent symptoms self-attributed to COVID-19. Data are mean ± standard error measure prevalence.

## Discussion

In this study, we included 30 consecutive patients seeking medical help for persistent symptoms (median 6 months) self-attributed to COVID-19. We identified two clinically comparable groups of long-COVID: those with and those without objective SARS-CoV-2 immunity. In 93% of patients, persistent symptoms had a cyclical pattern and were mostly represented by fatigue, thoracic oppression, myalgia, paresthesia and burning pain, which agrees with the literature (20,21). Dichotomizing patients by presence or not of anti-SARS-CoV-2 immunity resulted in only small differences in clinical presentation (**Figure 1B**). Indeed, both groups had high pain/fatigue indexes and limitations in several components of the SF-36 scale (**Supplemental Figures 1 and 3**).

Only half of the patient had objective cellular (IFN-γ ELISPOT-based) and humoral immunity for SARS-CoV-2 (**Figure 2B**). There are three possible (and non-exclusive) explanations for this result. First, some patients may have been infected with SARS-CoV-2, without detectable immunity. As already described (22), one of our patients with RT-PCR– proven SARS-CoV-2 infection had a negative SARS-CoV-2 serology result at several times (36, 85 and 140 days after initial symptoms) as well as on IFN-γ ELISPOT (after 140 days). Second, immunity may have developed in some patients but subsequently waned over time, although such reports are discordant (23,24). In our cohort, one patient with PCR/ELISPOT-confirmed infection showed IgG serology findings between days 133 and 251, both on anti-N (from equivocal to negative) and anti-S (from positive to equivocal) ELISA. Additionally, patients with cellular immunity might have negative serology, particularly anti-N IgG (**Figure 2A**). Finally, some patients may have presented a non-specific viral illness and subsequent symptoms, which were falsely attributed to SARS-CoV-2. In fact, the period of the first pandemic wave was highly anxiogenic and may have exacerbated pre-existing psychological conditions and induced a nocebo effect in some patients (25).

The high prevalence of probable anxiety and depression disorders (36.7% and 43.3% respectively) are striking. A cohort study in the United Kingdom revealed that the incidence of probable anxiety and depression disorder in the general population was 24% and 18.1% during the pandemic (26). A history of reported COVID-19 was associated with increased risk anxiety and depression. We also highlighted a high rate of probable PTSD (33.3%), which is similar to the 30.8% prevalence in a US study (27). PTSD reflects the hardship of the patient’s experience with COVID-19. The persistence of physical symptoms (related or not to COVID-19) is likely associated with the psychological burden of the pandemic, synergistically contributing to the emergence of “long-COVID”, and may explain the high prevalence of psychological disorders in our cohort.

Long-COVID is an ill-defined condition characterized by symptoms persisting for at least 4 to 12 weeks after SARS-CoV-2 infection, depending on the study (10,13). Excluding alternate diagnoses is crucial because objective evidence of previous COVID-19 may not be mandatory for the diagnosis of long-COVID (13,20). Deep phenotyping of our patients did not lead to alternative diagnoses in all patients. Long-COVID mainly represents patients with non-severe initial presentations (12), and its pathophysiology is likely multifactorial. For non-hospitalized long-COVID patients, physical deconditioning (28), psychological disorders (29), viral encephalitis (7), dysautonomia (30), and immunological abnormalities are plausible suspects and could co-exist. Indeed, auto-immunity has been demonstrated following COVID-19 (31,32). However, viral-induced autoimmunity is likely transient, as demonstrated by the negative repeat screening for antiphospholipid antibodies in 8 initially positive patients, as well as the lack of significant increase in cytokine/IFN-*α*2 levels and clinical/biological signs of autoimmune disease. Further studies will need to explore other potential mechanisms at play in such patients (*e*.*g*., dysautonomia, neurological studies).

This study has limitations. First, we included 30 consecutive patients, which may not allow to capture the complexity of long-COVID. However, this number allowed for a multimodal evaluation without missing data, and the consecutive recruitment limited potential selection bias. Given our study design, we cannot infer the prevalence of long-COVID, which has been evaluated at 10% in the United Kingdom (10). Only 9 patients had a RT-PCR test in our cohort in the 5 weeks after symptom onset, because RT-PCR tests were not available for mild diseases during the first epidemic wave in France. Finally, we cannot exclude that our design implied a selection bias of patients with psychological distress. However, our study allowed for accurately evaluating the patients seeking medical advice for persistent symptoms self-attributed to long-COVID and will therefore be of use for clinicians who may be under-prepared to answer these patients’ questions. Although no treatment has been approved for long-COVID, more than half of our patients reported a spontaneous and gradual improvement of symptoms over time, which provides a window of hope for these patients. To conclude, our study sheds light on the burden experienced by patients reporting long-term symptoms self-attributed to COVID-19, as well as some of the mechanisms at play. A better recognition and understanding of long-COVID will help healthcare providers care for these patients.

## Data Availability

Available on demand

## Authors contributions

MS, RF, JS, EC and LP recruited and evaluated clinically the patients. FG, CN and SFK conducted the virological evaluations. AM, AG and PV participated in the psychiatric evaluation. RAM conducted the psychological evaluation. SBT, RC, STA, MP, AB and SB conducted the immunological assays (cytokines). LA, JEG helped in the design of the study and critically reviewed the manuscript. MS, RF, FG, RFK and JS wrote the manuscript. MS, RF and FG are co-first authors. RFK and JS are co-last authors.

## Acknowledgments

We thank all patients who agreed to take part in the study. We thank Valle Meyer for her precious help in planning the study and patient communication. We also thank COVID-HUS for its support.

---

**Supplementary figure 1:**
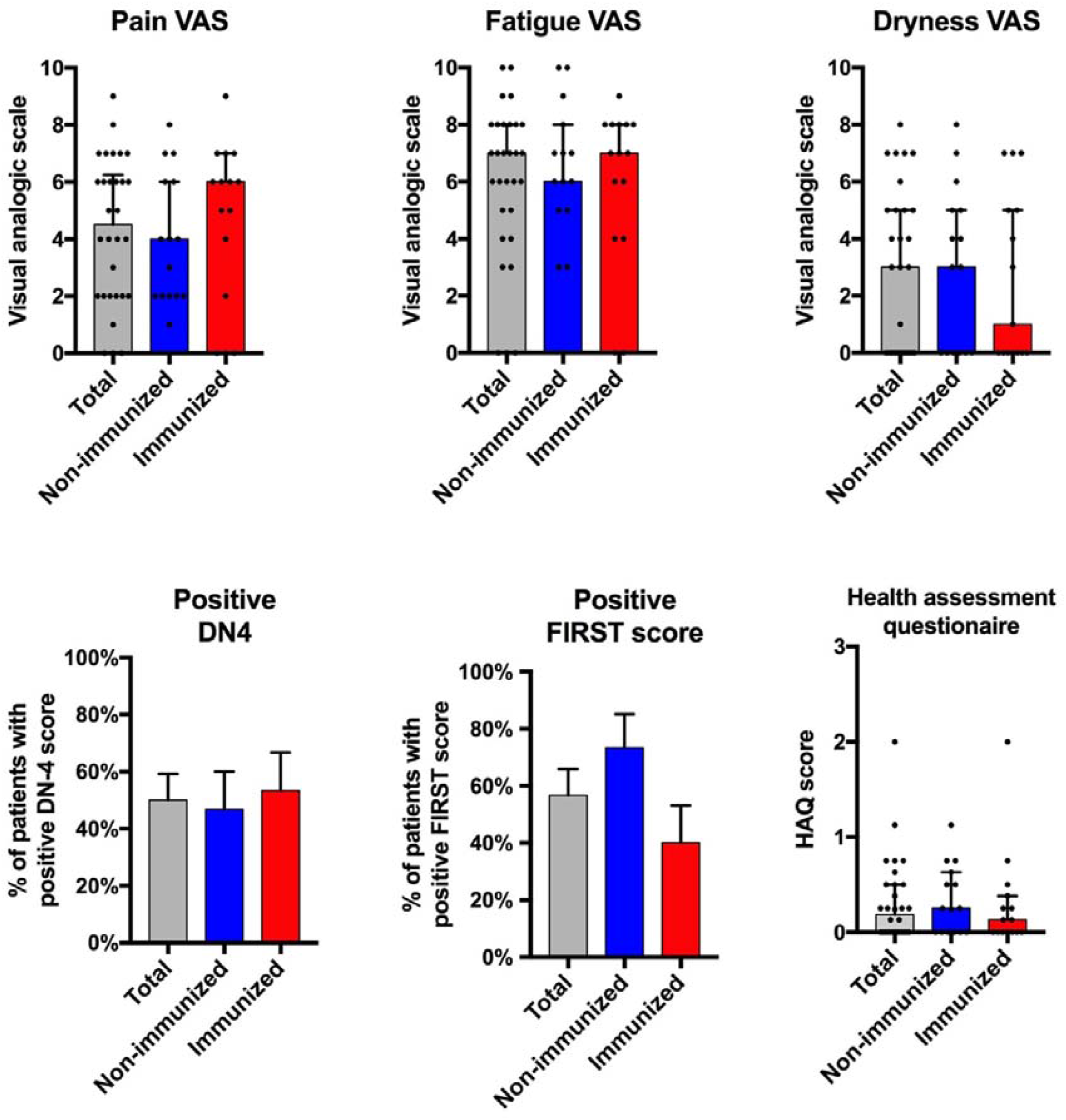
Comparison of pain, fatigue, dryness scores, DN4 and FiRST and Health Assessment Questionnaire (HAQ) scores in patients seeking medical help for persistent symptoms self-attributed to COVID-19. For continuous variables, data are median and interquartile range. For binary variables (DN4 and FiRST), data are mean± standard error measure prevalence of positivity. VAS, visual analog scale; FiRST, Fibromyalgia Rapid Screening Tool

**Supplementary figure 2:**
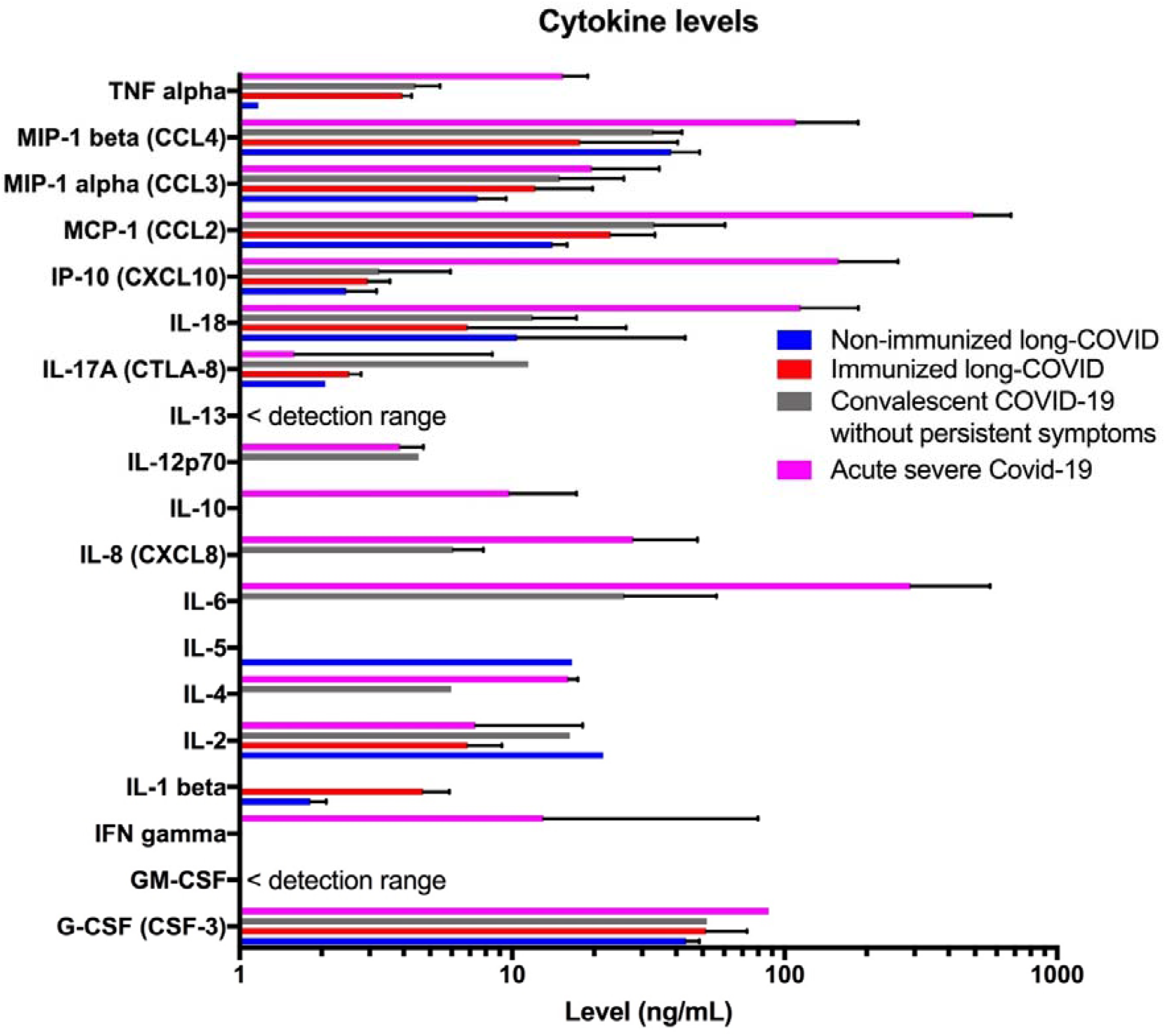
Cytokine levels in patients long-COVID patients compared to asymptomatic convalescent COVID-19 and acute severe COVID-19. Data are median and bars indicate interquartile range.

**Supplementary figure 3:**
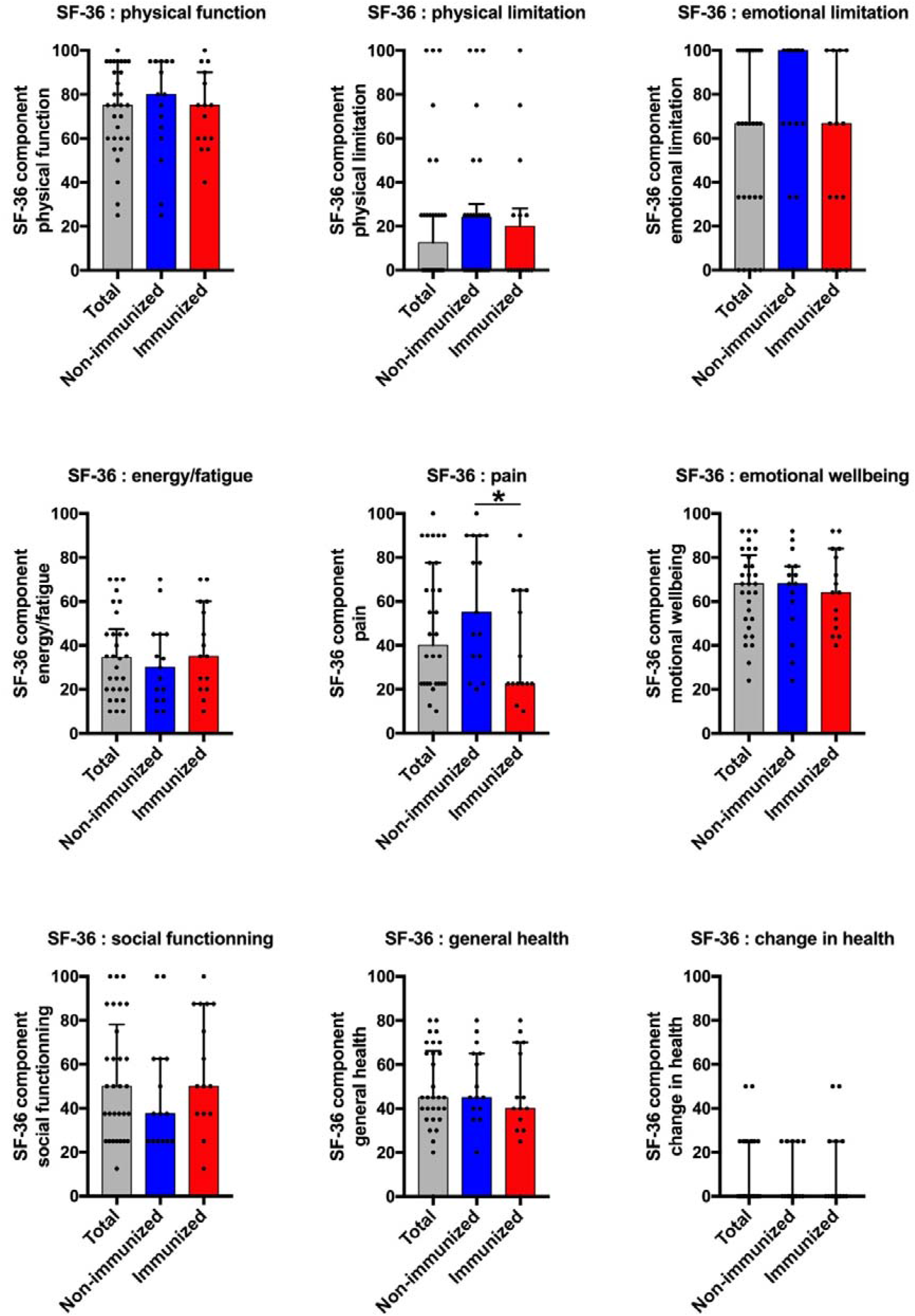
Components of the Medical Outcomes Study 36-item Short-form (SF-36) scale. Lower scores indicate worse health. Data are median and bars indicate interquartile range. *, p < 0.05 by Mann-Whitney test.

**Supplementary file 1: Systematic bloodwork of long-COVID patients**.

– Hemogram
– Ionogram, calcium, phosphorus, creatinine, urea, angiotensin-converting enzyme
– Creatine phosphokinase, troponin I
– Plasma protein electrophoresis and immunofixation
– Urine protein and creatinine
– TSH, 25 OH vitamin D, PTH
– TP, TCA, INR, D-dimers, fibrinogen
– Anti-nuclear antibodies (if ≥ 1/360: Extractable nuclear antibody and anti-double-stranded DNA were screened), rheumatoid factor, anti-citrullinated protein antibodies (ACPA using CCP2), ANCA, anti-cardiolipin, anti-beta-2 glycoprotein 1 antibodies and lupus anticoagulant.
– IgG, IgA, IgM levels
– C3-C4 and CH50
– HIV, HBV, HCV, CMV, EBV and Lyme (ELISA then western blot if positive) serology

